# Cohort Profile: Zhejiang Environmental and Birth Health Research Alliance (ZEBRA) Maternity Cohort

**DOI:** 10.1101/2023.02.21.23286173

**Authors:** Haitong Zhe Sun, Haiyang Tang, Qingyi Xiang, Siyuan Xu, Jing Fang, Haizhen Dai, Rui Shi, Yuxia Pan, Ting Luo, Hangbiao Jin, Chenyang Ji, Yuanchen Chen, Hengyi Liu, Meirong Zhao, Kung Tang, Yuming Guo, Wei Xu, Xiaoxia Bai, the Zhejiang Environmental and Birth Health Research Alliance (ZEBRA) collaborative group

## Abstract

The Zhejiang Environmental and Birth Health Research Alliance (ZEBRA) established a maternity cohort to investigate the relationship between perinatal abnormalities and various risk factors among the Chinese maternal population. The primary aim of ZEBRA is to explore the feasibility of early-stage risk prediction and the forecast of adverse perinatal symptoms and gestational outcomes. The cohort is ambidirectional, with a retrospective arm tracking 6,275 pregnant females enrolled between 2013 and 2016, and a prospective arm recruiting 112,414 participants since the baseline year of 2017. The current ZEBRA maternity cohort database comprises a diverse range of sociodemographic features, physiological characteristics, medical history, therapeutic interventions, and measurements of environmental exposures. Going forward, the cohort will continue to enrol a wider range of participants and collect an even more extensive array of features. ZEBRA is seeking collaborations with both national and international multi-cohort studies to contribute to the field of epidemiology, and provide valuable evidence-based insights for global maternal and child healthcare.

## Why was the ZEBRA maternity cohort set up?

Before the advent of modern medicine, childbirth carried an extremely high risk of maternal and perinatal mortality (the period beginning at the 20–28^th^ week of gestation and ending at 1–4 weeks after delivery). Fortunately, with advances in prenatal healthcare, the prevalence of maternal and perinatal mortalities has significantly reduced. However, gestational hypertension, postpartum haemorrhage, maternal infection, and other antepartum complications are still unneglectable health risks endangering the pregnant women. Under this circumstance, there is an urgent need to implement pragmatic measures to ensure the prolonged health of women during and after pregnancy.

Faced with the challenge of an ageing population in China, the central government has advocated the “three-child policy” since 2021 to optimise the population structure ^1^. This policy also stressed the importance of maintaining optimal maternal health. The probability of congenital disabilities, such as Down syndrome, neural tube malformation, and congenital heart disease, has been effectively controlled by popularising the new-generation high-sensitivity prenatal screening tools. These include the “double-index method” to investigate cardiac murmurs and transcutaneous oxygen saturation for the timely detection of congenital heart disease ^2^. However, the high occurrence of preterm births (e.g. ∼ 7.8% in China, ∼ 5.9% in Zhejiang) is still a concern ^3,4^. Preterm labour can increase the risks of multiple congenital disabilities, including low birth weight ^5^, immature organ development ^6^, cerebral palsy ^7^, and mental retardation ^8^.

Recent literature has highlighted the impact of environmental exposures on maternal health, encompassing a range of factors such as air pollutants ^9^, green spaces ^10^, and extreme temperatures ^11^. As a response to these hazards, the Zhejiang Environmental and Birth Health Research Alliance (ZEBRA) was established in 2022 under the leadership of the Health Commission of Zhejiang Province. ZEBRA aims to investigate the associations between endogenous physiological factors, exogenous environmental factors, and maternal health ^12^. Zhejiang Province was well-suited to lead this pioneering effort for several reasons. Firstly, it is one of the most socioeconomically developed provinces with low perinatal and neonatal mortality rates, allowing for a shift in research focus from individual-level clinical treatment to population-level risk prevention. Secondly, Zhejiang is a leader in cutting-edge interdisciplinary research with access to resources such as the Alibaba Cloud Intelligence Laboratory and DAMO Academy (Academy for Discovery, Adventure, Momentum and Outlook). The first phase of ZEBRA’s research (2022–2026) will focus on screening risk factors and exploring the possibility of early-stage prediction for perinatal abnormalities.

Perinatal abnormalities include a range of maternal symptoms such as gestational diabetes mellitus, gestational hypertension, pre-eclampsia ^13,14^, as well as preterm delivery (including spontaneous and indicated preterm birth) ^15^. The pathogenesis of perinatal abnormalities is complex and requires further exploration, making population-based cohort studies critical in safeguarding female health. Such studies can identify risk factors that cause perinatal abnormalities, such as insufficient nutrition intake during pregnancy ^15-17^, multiple gestations ^18,19^, pregnancy interval ^20^, workload ^21^, antepartum complications ^22^, psychological stress and depression ^23,24^, and environmental factors ^25-27^. The discoveries of the cohort study will be of significant public health significance for screening high-risk pregnant women, a major commitment made by ZEBRA upon its establishment.

Perinatal health protection necessitates collaborative efforts from both families and society, which is crucial for upholding gender equity. The health of women and children is indicative of a civilised society, promoting ongoing advancements in clinical obstetric medicine and public health research. ZEBRA seeks to address several key scientific inquiries. Firstly, are endogenous physiological characteristics and exogenous maternal environmental exposures correlated with perinatal abnormalities? Secondly, do endogenous and exogenous risk factors alter the impact of each other on the risk of perinatal abnormalities? Thirdly, can detectable longitudinal changes in physiological and biochemical markers of pregnant mothers serve as indicators of potential perinatal abnormalities? Fourthly, is it possible to predict the risks of pregnancy-related complications during the middle or even early stages of pregnancy using routine clinical tests? Additionally, is it feasible to achieve rapid risk prediction with easily accessible features that do not require cumbersome additional antenatal visits (e.g. features that can be collected on a smartphone), even among undereducated populations? During the process of epidemiological analysis, ZEBRA pledges to share periodic findings with the China Cohort Consortium ^28^, as well as cohort studies to improve strategies suitable for Chinese economic development and cultural practices.

### Who is in the cohort?

ZEBRA has been expanding a comprehensive maternity cohort since 2013, initiated in Zhejiang Province, with a goal to establish collaborations nationwide. Over the past few decades, the rate of hospital births in China has witnessed a significant surge, from 60.7% in 1996 to 99.7% in recent years ^29,30^. With a developed healthcare system, abundant public health resources, high popularity of multi-level medical units in towns and townships, and a strong emphasis on the health of the general population, the hospital birth rate in Zhejiang had already surpassed 99% in 2002 ^31^. This forms the social basis for ensuring maternal and neonatal health in the most comprehensive manner possible in Zhejiang Province.

ZEBRA enrols pregnant women and captures their pregnancy, delivery, and puerperium information, along with their neonates, as cohort participants. All pregnant women who choose to give birth at any hospital, adopting the same medical diagnostic criteria (to ensure homogeneity in information collection and inter-centre comparability), are eligible for inclusion in the ZEBRA Maternity Cohort, provided that they consent to share their relevant personal and non-private medical information. The construction and quality control of the ZEBRA multi-centre cohort is led by Hangzhou, the provincial capital city, in collaboration with other prefecture-level cities (Figure 1). The standardisation of the electronic medical system has propelled the ZEBRA Maternity Cohort into a completely new phase as of 2021. On December 9, 2021, the Zhejiang Health and Health Commission held a press conference on the comprehensive reform of mutual recognition and sharing of medical examination records. We are optimistic that more medical records of pregnant women will become eligible for cohort inclusion in future years, with further cross-province database sharing.

**Figure 1.**
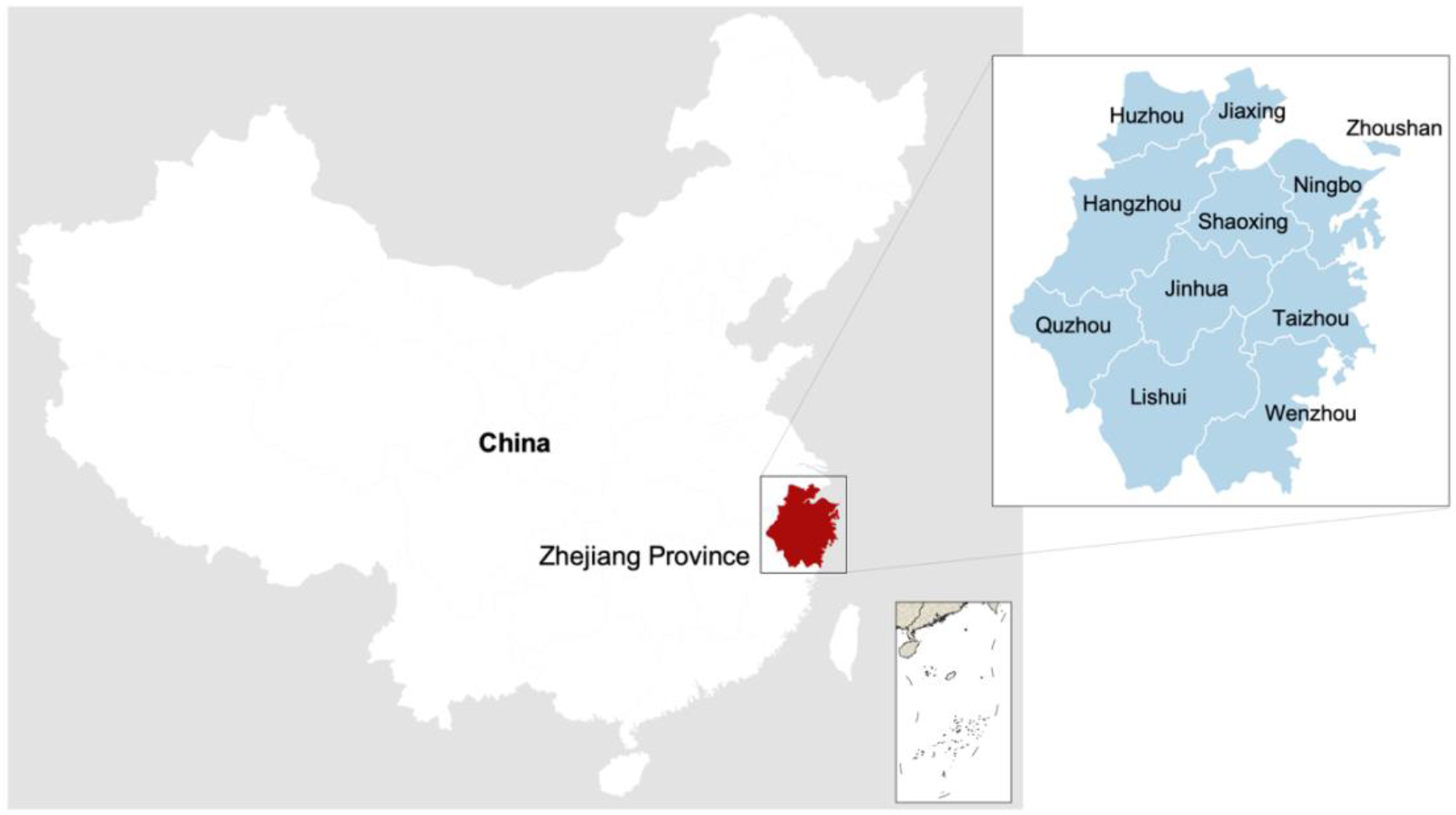
Location of Zhejiang Province and administrative map of prefecture-level cities.

Zhejiang Province, located in the Yangtze River Delta region on the east coast of China, spans 105,500 square kilometres (longitude 118°01’–123°10’ E, latitude 27°02’–31°11’ N). The province lies within the Humid Subtropical Climate Zone, characterized by significant monsoon alternation, moderate annual temperature, four distinct seasons, abundant sunshine, and rainfall. However, environmental problems, particularly air pollution, persist in Zhejiang Province. Despite effective reduction campaigns on several criteria air pollutants (i.e. PM_10_, sulphur dioxide, nitrogen dioxide, and carbon monoxide) in recent years ^32^, the excess rates of PM2.5 and ozone (O_3_) remain relatively high, at 6.8% and 4.9%, respectively, with peak O_3_ concentrations reaching 60–80 ppb ^33^. Such high O_3_ pollution poses population health hazards ^34^ and warrants future environmental epidemiological studies in Zhejiang Province.

The selection of Zhejiang as a priority province for the pioneering study of the large maternity cohort is justified by several reasons. Firstly, clinical treatment and primary care must take precedence over data-driven empirical studies, in keeping with the principle of humanism. In cases of limited resources, hospitals, medical schools, and medical research institutes must prioritise the prevention of maternal and neonatal deaths. While China has achieved significant reductions in maternal mortality rates through years of medical staff efforts, significant heterogeneity persists in the geographical distribution of mortality rates. Regions with low maternal mortality rates are concentrated in the east and southeast of China. In Zhejiang, the maternal and infant mortality rates were controlled within 7/100,000 and 5 ‰, respectively, maintaining the lowest levels in China. However, in southwest China, such as Yunnan, the maternal mortality rate was four times higher than that in Zhejiang Province in 2012. Therefore, for areas with high maternal mortality, the focus of medicine is still on the care of pregnant women and newborns. When the mortality rate is reduced to the forefront level of senior sister, research based on data will be carried out.

Secondly, Zhejiang’s critical role in the economic development of China also justifies its selection. In 2020, Zhejiang’s GDP reached 936.7 billion US dollars, accounting for 6.4% of the nation-level total GDP and ranking fourth among all provinces. Zhejiang’s economy even surpassed that of the Netherlands (909.1 billion US dollars), which ranks 17^th^ worldwide. This flourishing economy also promotes the popularisation of digital medicine, which provides a solid foundation for epidemiological studies, such as the ZEBRA Maternity Cohort and the Zhejiang Birth Cohort (ZBC).

Thirdly, geographical coverage and population representativeness were considered in the selection process. Beijing, Shanghai, and Zhejiang are the provincial regions with the lowest maternal and infant mortality rates in China, with the first two regions being municipalities directly under the Central Government. Beijing and Shanghai have attracted individuals with the highest education and economic status in China, with territorial areas of 16,411 and 6,340 square kilometres, respectively, which is far less than that of Zhejiang Province (101,800 square kilometres). Hence, Zhejiang Province is the ideal location to carry out digital maternal health public health research based on the current situation in China. The ZEBRA study also promises to share clinical experience and research findings with other regions of the country and even the world through the China Cohort Consortium (CCC), with the radiation driving less-developed regions to achieve geographical health justice.

## How often have the participants been followed up?

The ZEBRA maternity cohort is an open cohort that aims to recruit all pregnant women in Zhejiang Province and follow them throughout pregnancy until six months after delivery, at which point the follow-up will end if no abnormal health conditions are observed. During the pilot study conducted from 2013 to 2016, 6,275 participants were recruited by manually entering their information into the system due to the lack of a widely used interactive data archiving service in the medical system at that time. However, since 2017, Zhejiang Province has officially implemented systematic data management services. As of 31 December 2022, a total of 118,689 pregnant women have been enrolled in the ZEBRA maternity cohort, with the first antenatal visit recorded as the start and delivery (including miscarriage) as the end (Figure 2). All mid-term prenatal examinations of the same pregnant woman are consolidated and archived. Among them, 63,887 pregnant women underwent a 75 g oral glucose tolerance test (OGTT) at 24–28 weeks of gestation in hospitals implementing the same diagnostic standard as matched in the database. Future plans include continuing to follow up with the women and their newborns in the cohort, optimizing cohort sampling, increasing data acquisition channels, and ensuring the information security of cohort participants. Notably, ZEBRA is a cohort that is registered and tracked based on each pregnant woman’s prenatal visits, with no intervention in prenatal examination or treatment or additional financial burden imposed on the pregnant women ^3^.

**Figure 2.**
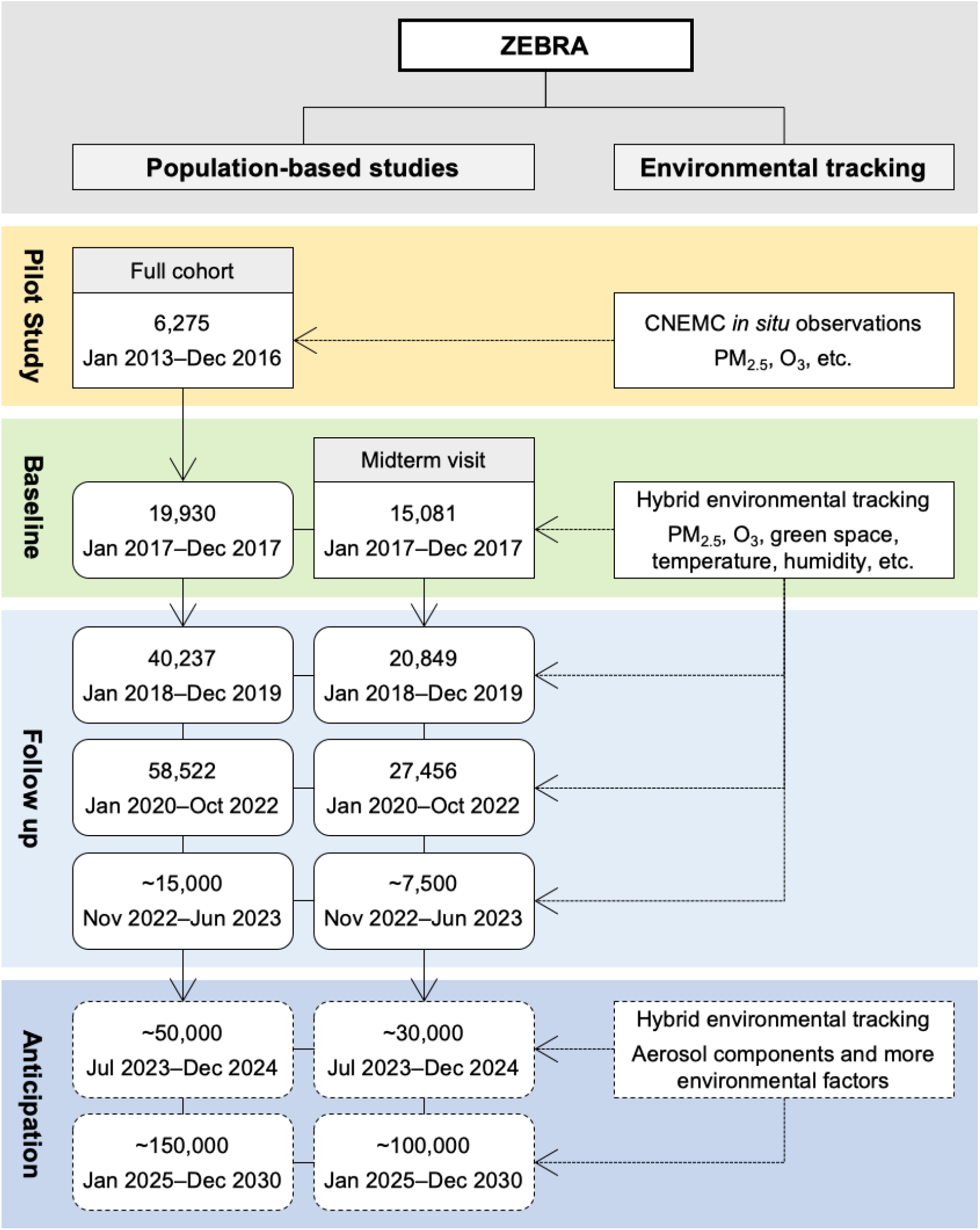
Flow diagram of pilot study, baseline recruitment, and follow-up for the Zhejiang Environmental and Birth health Research Alliance (ZEBRA) maternity cohort study.

The postpartum follow-up of the ZEBRA maternal cohort is an important aspect of the study, which continues until six months after delivery. The follow-up mainly includes the following procedures:

1. Within three days after delivery, the mothers undergo a set of fundamental physical examinations. These may include, but are not limited to, a blood routine test (which is decided independently by the obstetrician based on the condition of the pregnant woman), blood pressure measurement, and cardiac function evaluation. If the pregnant woman has pregnancy complications, targeted blood tests may be conducted.
2. We recommend that mothers undergo a hospital follow-up in the sixth week after delivery, which includes an assessment of weight recovery, blood pressure, and uterine involution. If the pregnant woman has pregnancy complications, targeted blood tests may be conducted.
3. We advise the mothers to report any abnormal health conditions within six months after delivery to the hospital by telephone. If the hospital has not received any such report, it will conduct a unified telephone follow-up in the sixth month, including an assessment of the mother’s blood pressure, weight, and breastfeeding. If the mother is healthy, the cohort follow-up ends, and the participants exit the cohort. However, if the mother has abnormal conditions or shows any signs of abnormal health, we will add a case-by-case follow-up for further clinical observation according to the situation.

The follow-up of neonates in the ZEBRA maternal cohort is conducted until six months after birth, and it mainly includes:

1. Documentation of birth weight and Apgar score at birth.
2. Clinical observation of jaundice, respiration, and feeding status within three days of birth.
3. Evaluation of weight and height development at 42 days after birth, and recording of other health abnormalities.
4. It is recommended that the mother report any abnormal health conditions of the neonate by telephone within six months after birth. If the hospital where the mother gave birth has not received any report, a unified telephone follow-up will be conducted in the sixth month to gather information.

## What has been measured?

Upon the inception of ZEBRA, a comprehensive investigation was conducted to gather sociodemographic characteristics, medical and disease history, as well as environmental exposure levels for the retrospective portion of the ambidirectional maternity cohort.

The sociodemographic characteristics comprise maternal age at delivery, ethnicity (Han or other minorities), occupation, educational attainment (graduate or above, undergraduate, college education, high school or below), place of residence (urban or rural), medical insurance type (New Rural Cooperative Medical Scheme, Urban Resident Basic Medical Insurance, and Urban Employee Basic Medical Insurance), and history of alcohol consumption and smoking, as summarised in Table 1.

**Table 1.** Statistics of sociodemographic features of ZEBRA maternity cohort participants, 2013–2021.

| Feature |  | Mean (SD) or Count (%) |
| --- | --- | --- |
| **Maternal Age at Delivery** |  | 30.9 (4.3) |
| **Height (metre)** |  | 1.6 (0.1) |
| **Pre-pregnancy BMI** |  | 26.4 (3.1) |
| **Ethnicity (Han)** |  | 116,671 (98.3%) |
| **Occupation** | Office staff | 94,494 (79.6%) |
|  | Worker | 13,929 (11.7%) |
|  | Professional and specialist | 2,879 (2.4%) |
|  | Farmer | 1,045 (0.9%) |
|  | Medical staff | 364 (0.3%) |
|  | Student | 157 (0.1%) |
|  | Others | 5,820 (4.9%) |
| **Education Attainment** | High school or below | 19,109 (16.1%) |
|  | College education | 23,263 (19.6%) |
|  | Undergraduate | 60,175 (50.7%) |
|  | Graduate or above | 16,142 (13.6%) |
| **Type of Residence** | Urban | 89,227 (75.2%) |
|  | Rural | 29,462 (24.8%) |
| **Type of Medical Insurance <sup>a</sup>** | NCMS | 8,730 (7.4%) |
|  | URBMI | 7,450 (6.3%) |
|  | UEBMI | 102,509 (86.4%) |
| **Alcohol and Smoking History** |  | 112 (0.1%) |

<sup>a</sup> Medical insurance include three types: New Rural Cooperative Medical Scheme (NCMS) for rural residents, Urban Resident Basic Medical Insurance (URBMI) for non-employed urban residents, and Urban Employee Basic Medical Insurance (UEBMI) for employees.

The electronic medical record system contains a total of 349 pre-pregnancy disease and medical treatment history items. Among these, 70 items have been identified as having an empirical association with reproductive health with prevalence rates higher than 0.05% (diseases with occurrence rates below 0.05% are defined as rare diseases, excluded for gestation-oriented epidemiological studies). Examples of such items include a history of cervical surgery, hepatitis B liver infection, anaemia, syphilis, intrahepatic cholestasis of pregnancy, and depression, among others. Table 2 provides an overview of the prevalence rates of 20 representative pre-pregnancy diseases and histories of surgeries, and Table 3 includes a list of 22 representative obstetric-relevant diagnostic indicators. These indicators include, but are not limited to, gestational weight gain, gestational day, history of live birth, stillbirth, miscarriage, menarche age, neonate birth weight, 1-minute and 5-minute Apgar scores. Full list of pre-pregnancy and gestational diagnostic items (a total of 54) was detailed in Supplementary Table 1 and 2, respectively. For each participant in the cohort, a comprehensive set of 60 physiological and biochemical indices were examined at the parturient period and during the 24–28^th^ gestational week oral glucose tolerance test (OGTT), as detailed in Supplementary Table 3. A summary of 28 particularly important indices is provided in Table 4.

**Table 2.**
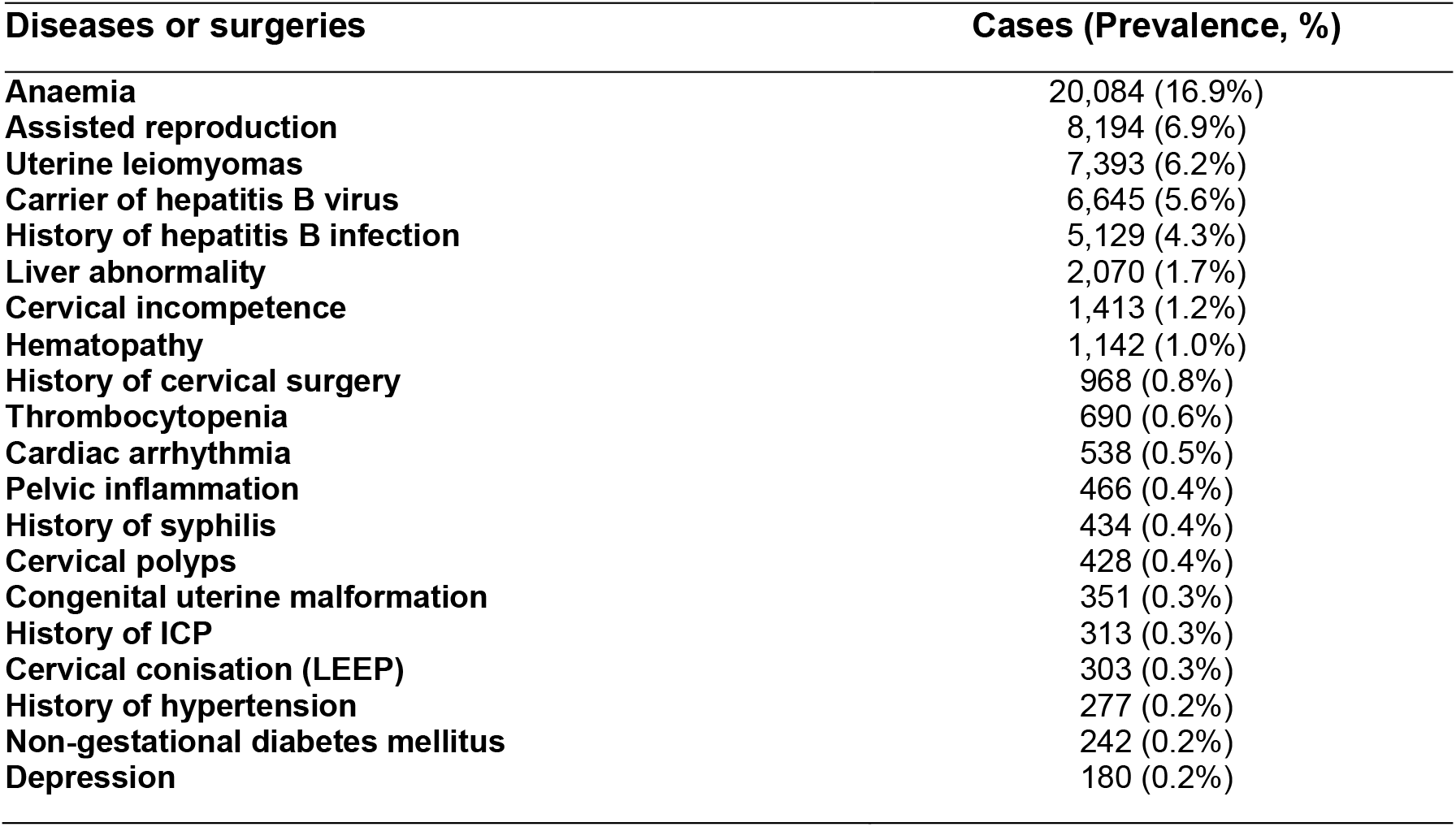
Cases and prevalence rates of 20 representative pre-pregnancy diseases and histories of surgeries. Items are listed in descending sequence of prevalence.

| Diseases or surgeries | Cases (Prevalence, %) |
| --- | --- |
| Anaemia | 20,084 (16.9%) |
| Assisted reproduction | 8,194 (6.9%) |
| Uterine leiomyomas | 7,393 (6.2%) |
| Carrier of hepatitis B virus | 6,645 (5.6%) |
| History of hepatitis B infection | 5,129 (4.3%) |
| Liver abnormality | 2,070 (1.7%) |
| Cervical incompetence | 1,413 (1.2%) |
| Hematopathy | 1,142 (1.0%) |
| History of cervical surgery | 968 (0.8%) |
| Thrombocytopenia | 690 (0.6%) |
| Cardiac arrhythmia | 538 (0.5%) |
| Pelvic inflammation | 466 (0.4%) |
| History of syphilis | 434 (0.4%) |
| Cervical polyps | 428 (0.4%) |
| Congenital uterine malformation | 351 (0.3%) |
| History of ICP | 313 (0.3%) |
| Cervical conisation (LEEP) | 303 (0.3%) |
| History of hypertension | 277 (0.2%) |
| Non-gestational diabetes mellitus | 242 (0.2%) |
| Depression | 180 (0.2%) |

**Table 3.**
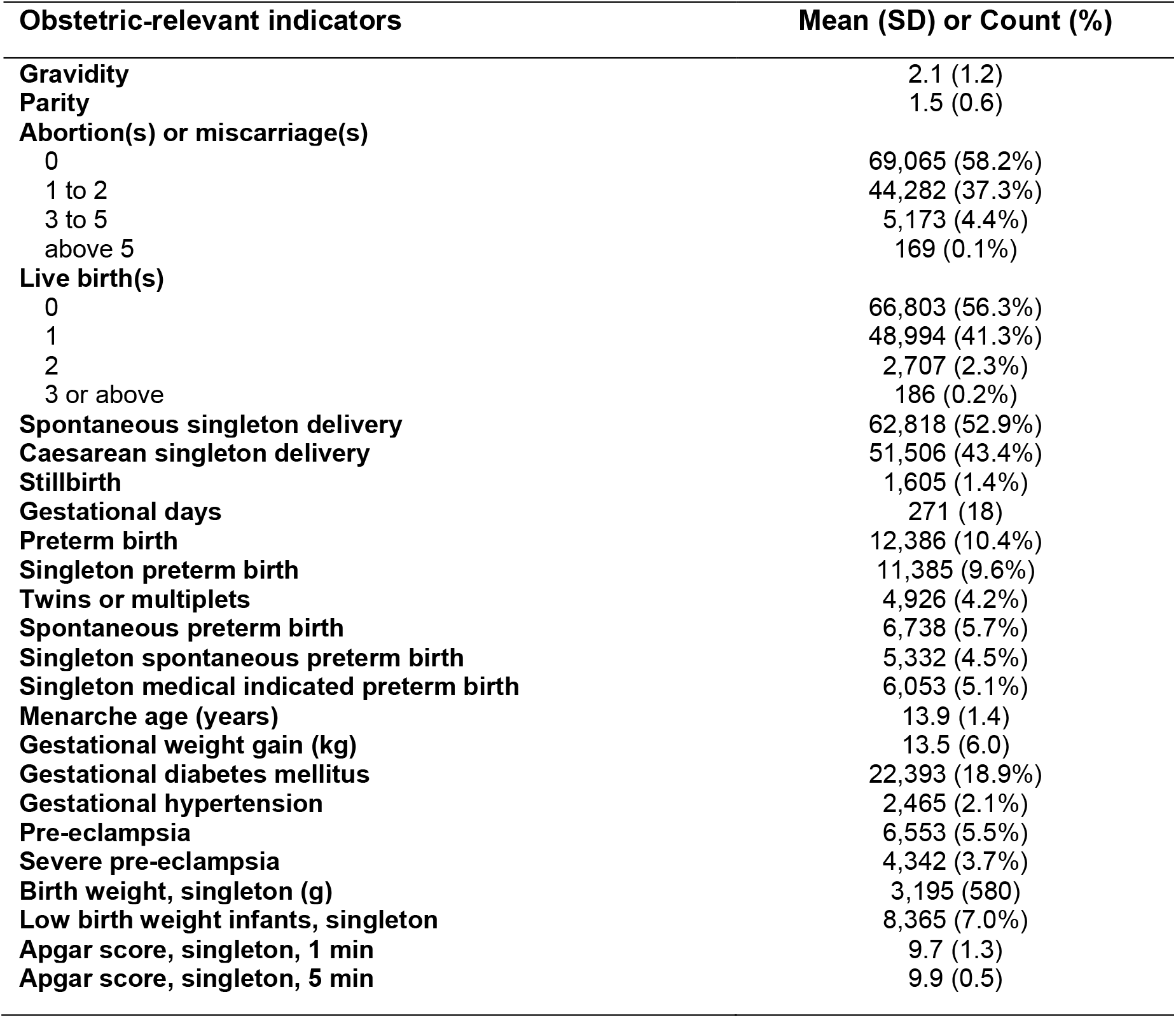
Statistics of obstetric-relevant diagnostic indicators.

**Table 4.**
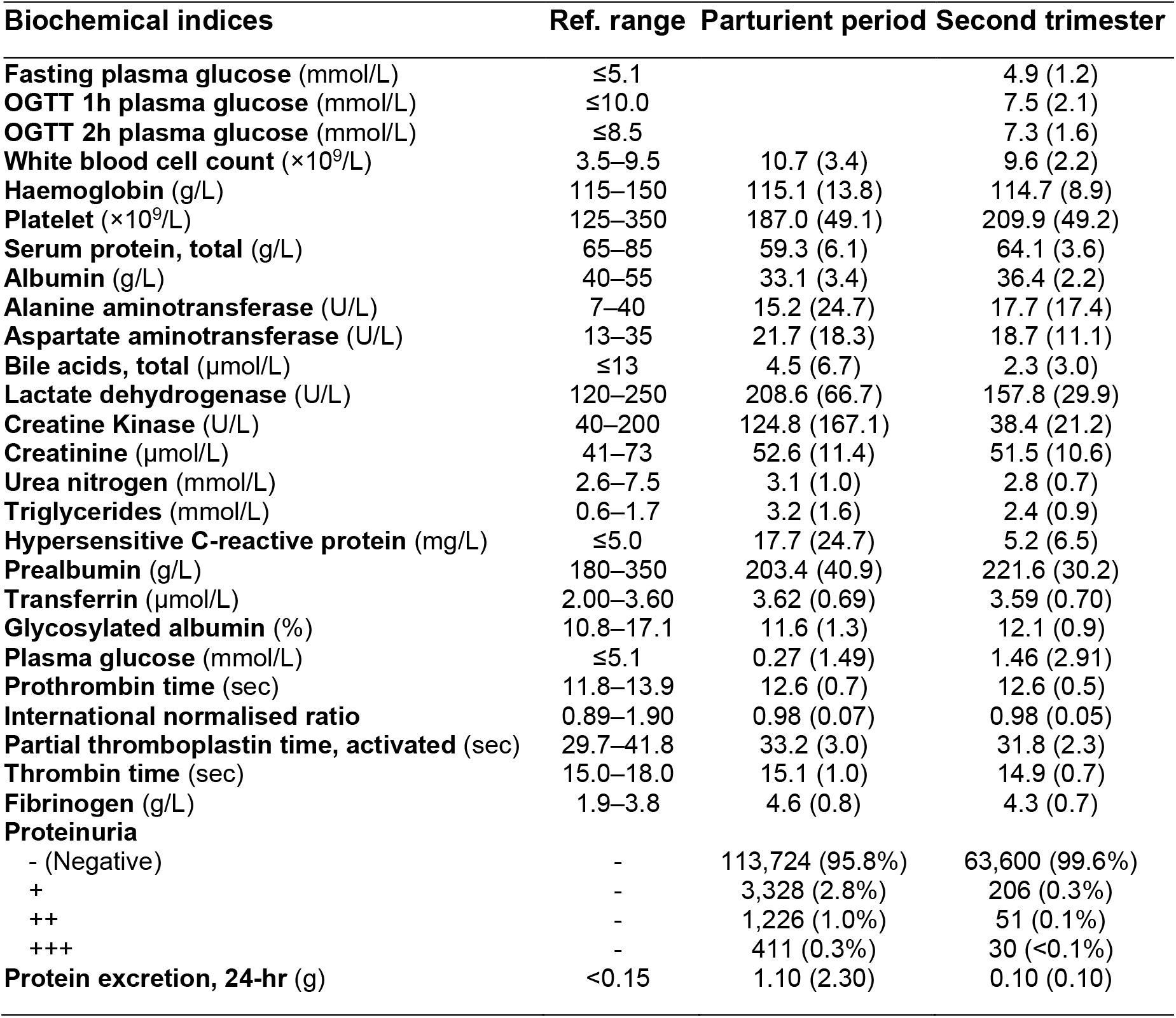
Representative biochemical indices at parturient period and oral glucose tolerance test (OGTT) in second trimester, 24–28^th^ gestational week.

| Biochemical indices | Ref. range | Parturient period | Second trimester |
| --- | --- | --- | --- |
| Fasting plasma glucose (mmol/L) | ≤5.1 |  | 4.9 (1.2) |
| OGTT 1h plasma glucose (mmol/L) | ≤10.0 |  | 7.5 (2.1) |
| OGTT 2h plasma glucose (mmol/L) | ≤8.5 |  | 7.3 (1.6) |
| White blood cell count (×10<sup>9</sup>/L) | 3.5–9.5 | 10.7 (3.4) | 9.6 (2.2) |
| Haemoglobin (g/L) | 115–150 | 115.1 (13.8) | 114.7 (8.9) |
| Platelet (×10<sup>9</sup>/L) | 125–350 | 187.0 (49.1) | 209.9 (49.2) |
| Serum protein, total (g/L) | 65–85 | 59.3 (6.1) | 64.1 (3.6) |
| Albumin (g/L) | 40–55 | 33.1 (3.4) | 36.4 (2.2) |
| Alanine aminotransferase (U/L) | 7–40 | 15.2 (24.7) | 17.7 (17.4) |
| Aspartate aminotransferase (U/L) | 13–35 | 21.7 (18.3) | 18.7 (11.1) |
| Bile acids, total (μmol/L) | ≤13 | 4.5 (6.7) | 2.3 (3.0) |
| Lactate dehydrogenase (U/L) | 120–250 | 208.6 (66.7) | 157.8 (29.9) |
| Creatine Kinase (U/L) | 40–200 | 124.8 (167.1) | 38.4 (21.2) |
| Creatinine (μmol/L) | 41–73 | 52.6 (11.4) | 51.5 (10.6) |
| Urea nitrogen (mmol/L) | 2.6–7.5 | 3.1 (1.0) | 2.8 (0.7) |
| Triglycerides (mmol/L) | 0.6–1.7 | 3.2 (1.6) | 2.4 (0.9) |
| Hypersensitive C-reactive protein (mg/L) | ≤5.0 | 17.7 (24.7) | 5.2 (6.5) |
| Prealbumin (g/L) | 180–350 | 203.4 (40.9) | 221.6 (30.2) |
| Transferrin (μmol/L) | 2.00–3.60 | 3.62 (0.69) | 3.59 (0.70) |
| Glycosylated albumin (%) | 10.8–17.1 | 11.6 (1.3) | 12.1 (0.9) |
| Plasma glucose (mmol/L) | ≤5.1 | 0.27 (1.49) | 1.46 (2.91) |
| Prothrombin time (sec) | 11.8–13.9 | 12.6 (0.7) | 12.6 (0.5) |
| International normalised ratio | 0.89–1.90 | 0.98 (0.07) | 0.98 (0.05) |
| Partial thromboplastin time, activated (sec) | 29.7–41.8 | 33.2 (3.0) | 31.8 (2.3) |
| Thrombin time (sec) | 15.0–18.0 | 15.1 (1.0) | 14.9 (0.7) |
| Fibrinogen (g/L) | 1.9–3.8 | 4.6 (0.8) | 4.3 (0.7) |
| Proteinuria |  |  |  |
| - (Negative) | - | 113,724 (95.8%) | 63,600 (99.6%) |
| + | - | 3,328 (2.8%) | 206 (0.3%) |
| ++ | - | 1,226 (1.0%) | 51 (0.1%) |
| +++ | - | 411 (0.3%) | 30 (<0.1%) |
| Protein excretion, 24-hr (g) | <0.15 | 1.10 (2.30) | 0.10 (0.10) |

The establishment of a high-resolution spatiotemporal database has enabled the tracking of individual-level exposures to criteria air pollutants, such as particulate matters (i.e. PM2.5) ^35,36^ and ozone ^33,37,38^, as well as green space ^39^, humidity, and extreme temperatures ^40^ through geospatial projection based on residential location. In the future, with the full operation of the cloud medicine platform, we propose to collect lifestyle features, such as dietary habits, physical exercise, sleeping patterns, and mental conditions, as well as behavioural changes for climate adaptation, and history of medicine and supplement use through smartphone applications. Wearable devices combining chemical and optical probes, pedometers, and GPS are under development to enable high-resolution time-series tracking of personal air pollution exposure, which is scheduled to begin implementation in 2024. For participants willing to provide further biological samples, exposure levels to heavy metals and organic chemical pollutants will also be determined, for example, in placenta samples. Due to limitations in detection techniques and public reluctance to share genetic information, the ZEBRA cohort does not plan to collect gene-level information from participants at this time.

## What has it found? Key findings and publications

ZEBRA has played a critical role in addressing research gaps related to endogenous (such as sociodemographic and physiological characteristics) and exogenous (such as medical history and environmental exposure) risk factors associated with maternal, perinatal, childbirth, and neonatal abnormalities among Chinese females. As of now, ZEBRA has published more than 20 peer-reviewed research articles, reports, reviews, and news pieces. Previous research conducted by ZEBRA primarily involves microscale laboratory experiments, as well as data-driven macroscale population-based epidemiological studies. For the purposes of this summary, we have selected publications that are of significant clinical or public health importance.

### Uterine rupture risk factor identification

We identified 19 cases of uterine rupture among pregnant women who had not previously undergone caesarean section between 1992 and 2017. We collected demographic and sociological characteristics, clinical features, and details on the location and extent of the rupture (i.e. complete or incomplete), as well as pregnancy complications and childbirth risk factors (e.g. prolonged labour, dystocia, induced labour, use of oxytocin, etc.). We found that a history of curettage and multiple pregnancy were associated with complete uterine rupture, which was more likely to occur in the fundus of the uterus and at a smaller gestational age, leading to adverse pregnancy outcomes. Neonates with incomplete uterine rupture had higher Apgar scores and lower perinatal mortality ^41^. While findings from these rare cases may be used for clinical guidance, the limited sample size restricts generalisation, highlighting the need for more cases from the ZEBRA maternity cohort in the future to support these findings.

### Neonatal organic pollutant exposure

Currently, research into the pregnancy exposure characteristics and transplacental metastasis of isoforms of per- and polyfluoroalkyl compounds (PFASs) is limited. Such evidence not only advances our comprehension of the mechanism of transplacental transfer of PFASs, but also provides guidance for controlling the production process associated with particular PFASs to lessen occupational and daily exposure. To enhance our understanding of the transplacental and breastfeeding transfer of PFASs, and provide data that supports neonatal health risk assessment, we gathered mother-child paired maternal serum, cord serum, breast milk, or placenta samples from three Chinese cities - Hangzhou, Wuhan, and Mianyang - to capture geographical variability for the determination of various PFAS species’ concentrations. The mean concentration of ∑ PFAS in maternal serum followed the order of Mianyang (4.44 ng/mL) < Wuhan (9.88 ng/mL) < Hangzhou (19.72 ng/mL) ^42^. Perfluorooctanoic acid (PFOA), perfluorooctanesulfonic acid (PFOS), and 6:2 chlorinated polyfluorinated ether sulfonates (6:2 Cl-PFESA) were the most prevalent PFASs found in serum ^43,44^. Linear and branched PFOS and PFOA can be effectively transferred across the placenta, with exposure levels ranked in the order of maternal serum > cord serum > placenta ^45,46^. Moreover, we have critically summarised the current knowledge on the inner exposure status and mother-to-infant transfer characteristics of PFASs, drawing on hundreds of studies published from 2000 to date, in order to better comprehend this field ^47,48^.

In addition, we collected paired serum and cerebrospinal fluid samples from 49 and 22 neonates, respectively, to determine the concentrations of PFASs in these biological samples. Our findings showed positive concentrations of PFASs in the cerebrospinal fluid, leading us to conclude that both PFASs and emerging PFAS substitutes have the ability to penetrate the developing blood-cerebrospinal fluid barrier (BCSFB), thus posing a potential risk to the healthy development of newborns ^49^.

### Air pollution exposure and preterm birth risks

Air pollution has been identified as a critical risk factor for maternal health hazards such as miscarriage, preterm birth, and stillbirths, as reported in existing literature ^50-52^. In this study, we recruited 6789 pregnant mothers with stratified randomisation from 2013 to 2017 in ZEBRA, with a preterm birth prevalence of 5.5% ^3^. We monitored individual-level exposure to criteria air pollutants and estimated that the risk of preterm birth would increase by 13% (95% confidence interval, CI: 3–25%) and 12% (95% CI: 5–19%) with every 10 μg/m^3^ additional exposure to PM_2.5_ and O_3_, respectively. Additionally, we found that low body mass index (BMI), multiple gravidity times, and gestational hypertension also contributed to the risk of preterm birth occurrence.

### Mother-to-child intergenerational transmission

In order to investigate the potential transmission of Hepatitis B Virus (HBV) from mother to infant, we obtained placental tissues from three groups of women: 30 healthy pregnant women who were not infected with HBV (HBV^-/-^), 30 HBV-positive women who did not infect their neonates (HBV^+/-^), and 30 HBV-positive women who did infect their neonates (HBV^+/+^). Additionally, peripheral blood samples were collected from the femoral vein of six HBV^+/+^ and six HBV^+/-^ neonates. Serum protein expression profiles of HBV were analysed by proteomics, and the expression of proteins related to intrauterine HBV transmission was further confirmed through immunohistochemistry and PCR techniques in both serum samples and placental tissues. After screening and identifying 35 differential proteins, we found that high expression levels of S100-family proteins and exocytosis mediated by the AnxA2-S100A10 complex were associated with intrauterine HBV transmission through the placenta ^53,54^.

The intergenerational effects of maternal antibiotic abuse on mother-child health have been a subject of concern among public health researchers, given the evidence of potential hazards to the healthy growth and development of children. Notably, the damage to gut bacteria is a significant outcome of antibiotic exposure ^55^. In light of this, we collected breast milk and faecal samples from 25 mothers who received either cefuroxime (CXM) or CXM-cefoxitin (CFX) combined treatments, with faecal samples also obtained from their infants in 2020. Of the 25 participants, nine received no antibiotic treatment, 13 received CXM treatment, and three received CXM + CFX treatment. At a 6-month postpartum follow-up, we collected faecal samples from five infants whose mothers had undergone antibiotic treatments. We analysed the microbiota in both breast milk and faecal samples to investigate the undesirable effects of antibiotics on the microbiota. Furthermore, we compared the relative abundance of antibiotic resistance genes (ARGs) in the infant gut microbiota to investigate their transfer. While antibiotic treatments had no influence on the microbiota of breast milk, they did disturb the gut microbiota in neonates. The abundance of ARGs in the infant gut microbiota showed a declining trend in the antibiotic-treated groups but significantly increased after 6 months of recovery ^56^.

### Out-of-balance eating quality during COVID-19 pandemic

A cohort of 3689 pregnant women from 19 provinces or municipalities in China were recruited to participate in an online questionnaire survey from 23 January to 29 February 2020, the period classified as the “emergency phase” of COVID-19 epidemic prevention and control in the country. The survey collected demographic information, food intake frequency, and health-related behavioural data of the participants. The study revealed that the overall diet quality of pregnant women was moderately unbalanced during the aforementioned period. The consumption of vegetables, fruits, livestock or poultry meat, dairy products, and nuts were all decreased. Notably, overeating of cereals and potatoes was a major concern in many regions due to restricted access to food supply caused by the lockdown measures ^57^.

### Gestational effects from severe diseases

Gestational trophoblastic neoplasia (GTN) is a specific tumour that affects pregnant women, originating from the malignant transformation of trophoblastic cells in placental villi during pregnancy. Chemotherapy has been recognised as an effective method to treat GTN, although concerns have been raised regarding its potential impact on pregnancy outcomes. In order to investigate this potential effect, we recruited 319 pregnant patients diagnosed with low-risk GTN in 2018 from the ZEBRA cohort. The patients were divided into two groups according to the occurrence of severe myelosuppression. We found that the occurrence of severe myelosuppression did not have an impact on the chemotherapy effect of methotrexate or the pregnancy outcomes ^58^.

Gestational diabetes mellitus (GDM) is a crucial issue that has an impact on pregnancy outcomes and the long-term health of pregnant women. To investigate the feasibility of an early clinical diagnosis of GDM, we recruited all pregnant women diagnosed with GDM from the ZEBRA maternity cohort in 2019 and randomly selected healthy pregnant women with normal glucose tolerance as the control group. We used qRT-PCR to detect the expression level of the gene miR-520h in serum samples, and analysed the association between fasting blood glucose (FBG) level and miR-520h expression. Our results revealed that the expression level of miR-520h was up-regulated in the serum of GDM patients. We concluded that miR-520h could serve as a potential biomarker for the earlier-stage diagnosis of GDM ^59^.

ZEBRA has directed attention towards understanding the pathological progression of endometrial carcinoma (EC). To this end, we obtained endometrial samples from 345 ZEBRA participants between 2006 and 2013, consisting of 55 normal endometrium (NE), 27 atypical hyperplasia (AH), and 263 Type-I EC. Immunohistochemical staining was performed to investigate the correlation between the expression of the TWIST1 chromosome and survival, as well as clinicopathological characteristics. Our findings indicate that TWIST1 expression gradually increased from NE to AH, implying that heightened TWIST1 expression is a significant indicator of the progression from hyperplasia to AH and eventually malignant tumour. Furthermore, we observed that high TWIST1 expression in Type-I EC patients suggested an increased invasive ability and a greater risk of distant metastasis of the tumour 60.

At ZEBRA cohort, maternal serum samples were used to participate in the validation of fluorescent probes for the diagnosis of intrahepatic cholestasis of pregnancy (ICP). This study supported the use of a novel diagnostic tool for effective detection of ICP, and promoted the development of new biochemical markers that can better characterize maternal liver function 61.

## What are the main strengths and weaknesses of the study?

There are four major strengths of research conducted on ZEBRA maternity cohort. Firstly, to our best knowledge, the ZEBRA maternity cohort represents the first comprehensive pregnancy and childbirth cohort study in China that incorporates various intrinsic factors such as physiological and biochemical characteristics, complete medical histories, and extrinsic factors like environmental exposure, to minimise the bias in risk estimation caused by solely investigating exogenous risk factors as in previous studies. Secondly, in addition to comprehensively evaluating a range of potential risk factors, ZEBRA aims to explore the interaction effects between endogenous and exogenous factors to identify high-risk pregnant women. Thirdly, with more than 0.1 million pregnant women in the cohort, and close to 300 tracked factors for each participant, the vast dataset can be employed to develop risk prediction models for early-stage identification of abnormal perinatal outcomes. Fourthly, since 2022, the popularisation of “Cloud Healthcare” has enabled the amalgamation of hospital diagnostic records and self-reported information from smartphone applications to collect health information in a more comprehensive and reliable manner while ensuring the privacy and security of personal information of the cohort participants.

The present stage of the ZEBRA maternity cohort study still has several main limitations. Firstly, during the pilot study conducted during 2013–2017, the estimated relative risk values between preterm birth and maternal exposure to air pollutants were highly uncertain due to the small sample size and the lack of effective adjustment for multi-pollutant cross-confounding effects. The risk estimations will be updated in subsequent comprehensive epidemiological studies involving a wider cohort (i.e. from ∼ 6000 to over 0.1 million participants). Secondly, while the current data structure used for analysis categorises populations based on physiological and biochemical indicators and medical history, genetic factors are theoretically the primary method for distinguishing endogenous differences. However, due to ethical requirements, national-level genetic research control, limitations in efficient detection technology, and lower willingness of pregnant women to share genetic information, genetic-level cohort studies are not scheduled to commence before 2025. Lastly, the current ZEBRA cohort study considers relatively few variables related to behavioural characteristics, which are theoretically only obtainable through self-report by participants and thus often have lower credibility. The “Cloud Healthcare” platform is anticipated to enhance the client-based questionnaire survey through artificial intelligence-driven optimisation in the next two years: improvements in the frequency and capacity of data collection, and the implementation of double verification are among the goals for achieving more reliable self-reported indicators by 2025.

ZEBRA has pledged to provide evidence-based research conclusions on maternal health for the Chinese population, considering that the majority of high-quality cohort studies have been established in Western countries. ZEBRA aims to set an example in the standardised management of electronic medical records for mother-child pairs at a provincial and national level. Optimised models for screening risk prediction and identification of risk factors using data mining techniques can be promptly applied to newly admitted pregnant women. This approach will improve maternal health risk assessment and update dynamically with the inclusion of cohort participants. Moreover, ZEBRA will take on the responsibility of disseminating maternal health research to the general public, hoping to reduce risks as early as possible.

At present, numerous factors are affecting birth rates among young people. However, gynaecologists, physicians, nurses, and medical students are still dedicated to providing the most professional and comprehensive life and healthcare services for women who wish to give birth. The medical and research community is committed to leaving no one behind. By establishing ZEBRA, Zhejiang aims to contribute to the achievement of the “Healthy China 2030” vision and the United Nations’ Sustainable Development Goals, including SDG3 in maternal and child health, by publishing high-quality research reports and providing epidemiological evidence for global maternal and child healthcare.

## Can I get hold of the data? Where can I find out more?

ZEBRA is a valuable resource for the research community, offering a wide range and depth of clinical diagnostic information and individual-level tracking of environmental exposure (e.g. PM_2.5_, O_3_, green space and abnormal temperature). While the study database is not publicly available, ZEBRA warmly welcomes potential collaborations, and such requests are considered on a case-by-case basis. Researchers seeking access to the dataset are required to obtain approval from the designated ZEBRA research proposal review committee, comprising the School of Medicine at Zhejiang University and the Maternal and Child Health Division of the Health Commission of Zhejiang Province. International researchers seeking additional information regarding collaboration and data access are encouraged to contact the corresponding authors, Dr. Xiaoxia Bai and Dr. Haitong Z. Sun, and to specify their requests.

## Cohort profile in a nutshell

### Zhejiang Environmental and Birth Health Research Alliance (ZEBRA) maternity cohort

- The ZEBRA maternity cohort was formulated to examine the links between several categories of perinatal complications and various risk factors in the Chinese maternal population. A paralleled aim is to determine the feasibility of screening and forecasting potential risks during the early stages of pregnancy.

- The ZEBRA maternity cohort is a unique and comprehensive cohort that operates on an ambidirectional basis. The retrospective arm of the cohort tracks 6,275 pregnant females who were enrolled between 2013 and 2016.

- The prospective arm has already recruited 112,414 participants since the baseline year of 2017. Going forward, the cohort will continue to enrol a wider range of participants and collect an even more extensive array of features.

- The dataset of the ZEBRA maternity cohort encompasses a diverse array of sociodemographic features, physiological characteristics, medical history, therapeutic interventions, and measurements of environmental exposures.

- ZEBRA seeks to establish collaborations with both national and international multi-cohort studies, with the ultimate goal of contributing to the field of epidemiology and providing valuable evidence-based insights for global maternal and child healthcare.

## Data Availability

While the study database is not publicly available, ZEBRA warmly welcomes potential collaborations, and such requests are considered on a case-by-case basis. Researchers seeking access to the dataset are required to obtain approval from the designated ZEBRA research proposal review committee, comprising the School of Medicine at Zhejiang University and the Maternal and Child Health Division of the Health Commission of Zhejiang Province. International researchers seeking additional information regarding collaboration and data access are encouraged to contact the corresponding authors, Dr. Xiaoxia Bai and Dr. Haitong Z. Sun, and to specify their requests.

## Ethics approval

The study obtained written informed consent from all participants, and the research involves the use of de-identified personal information for scientific purposes. The ZEBRA cohort study is approved by the ethics committee of the Women’s Hospital, School of Medicine, Zhejiang University (IRB-20220189-R).

## Acknowledgments

The authors wish to extend their appreciation to several entities for their contributions in the establishment of ZEBRA. Special appreciations to Professor Weiguo Lu, the chief director of the Key Laboratory of Women’s Reproductive Health, located in Hangzhou, Zhejiang Province. Specifically, (1) the Department of Maternal Healthcare at Women’s Hospital, School of Medicine, Zhejiang University, and (2) the Maternal and Child Health Division at the Health Commission of Zhejiang Province, for providing comprehensive supports. In addition, (3) the TAP (Tracking Air Pollution in China) team at Tsinghua University, (4) the Institute of Reproductive and Child Health, National Health Commission Key Laboratory of Reproductive Health, and the Department of Epidemiology and Biostatistics at the School of Public Health, Peking University Health Science Centre, (5) the School of Atmospheric Sciences at Nanjing University, (6) the Vanke School of Public Health at Tsinghua University, and (7) the School of Public Health and Preventive Medicine at Monash University (Australia) are also acknowledged for their technical assistance. Finally, the authors express gratitude to all ZEBRA cohort participants, as well as the coordinators, administrative and technical staff, and participant interviewers who have made valuable contributions to the cohort study.

