## Supplementary Materials for "Cohort Profile: Zhejiang Environmental and Birth Health Research Alliance (ZEBRA) Maternity Cohort"

**Supplementary Table 1. Full list of cases and prevalence rates of pre-pregnancy diseases and histories of surgeries.** Items are listed in descending sequence of prevalence.

| Disease history | Cases (Prevalence, %) |
|---|---|
| Anaemia | 20,084 (16.9%) |
| Assisted reproduction | 8,194 (6.9%) |
| Uterine leiomyomas | 7,393 (6.2%) |
| Carrier of hepatitis B virus | 6,645 (5.6%) |
| History of hepatitis B infection | 5,129 (4.3%) |
| Mesosalpinx cyst | 2,975 (2.5%) |
| Streptococcus carrier | 2,813 (2.4%) |
| Thrombophilia | 2,139 (1.8%) |
| Ovarian cyst | 2,107 (1.8%) |
| Liver abnormality | 2,070 (1.7%) |
| Immune system diseases | 1,950 (1.6%) |
| Cervical incompetence | 1,413 (1.2%) |
| Hematopathy | 1,142 (1.0%) |
| Hashimoto thyroiditis | 1,049 (0.9%) |
| Placenta implantation (accreta/increta/percreta) | 1,023 (0.9%) |
| Antiphospholipid syndrome | 974 (0.8%) |
| History of cervical surgery | 968 (0.8%) |
| Mycoplasma infection | 933 (0.8%) |
| Cholecystolithiasis and cholecystitis | 881 (0.7%) |
| Endometriosis | 790 (0.7%) |
| History of malignant neoplasm of thyroid gland | 692 (0.6%) |
| Thrombocytopenia | 690 (0.6%) |
| Candida vaginitis | 680 (0.6%) |
| Septate uterus | 648 (0.5%) |
| Membranitis | 622 (0.5%) |
| Vaginitis | 589 (0.5%) |
| Cardiac arrhythmia | 538 (0.5%) |
| History of hypothyroidism | 510 (0.4%) |
| Abnormal umbilical arterial flow | 484 (0.4%) |
| Single umbilical cord artery | 468 (0.4%) |
| Pelvic inflammation | 466 (0.4%) |
| Uterine rupture | 459 (0.4%) |
| History of syphilis | 434 (0.4%) |
| Cervical polyps | 428 (0.4%) |
| Adenomyosis | 428 (0.4%) |
| Thalassemia | 416 (0.4%) |
| Hyperthyroidism | 403 (0.3%) |
| Polycystic ovary syndrome | 403 (0.3%) |
| History of thyroid surgery | 403 (0.3%) |
| Total thyroidectomy associate | 355 (0.3%) |
| History of gallbladder disease | 355 (0.3%) |
| Congenital uterine malformation | 351 (0.3%) |
| History of ICP | 313 (0.3%) |
| Cervical conisation (LEEP) | 303 (0.3%) |
| Nephropathy and nephritis | 291 (0.2%) |
| History of hypertension | 277 (0.2%) |
| Subtotal thyroidectomy | 250 (0.2%) |
| Non-gestational diabetes mellitus | 242 (0.2%) |
| Cardiac insufficiency | 233 (0.2%) |
| History of Hashimoto's thyroiditis | 228 (0.2%) |
| Amniotic fluid pollution | 201 (0.2%) |
| History of Preeclampsia | 182 (0.2%) |
| Depression | 180 (0.2%) |
| Congenital heart disease | 174 (0.1%) |
| Bacterial vaginitis | 165 (0.1%) |
| Urolithiasis | 159 (0.1%) |
| Didelphic uterus | 159 (0.1%) |
| Ankylosing spondylitis | 157 (0.1%) |
| Intrauterine death | 141 (0.1%) |
| Hellp's syndrome | 140 (0.1%) |
| Antiphospholipid syndrome | 125 (0.1%) |
| Total thyroidectomy | 105 (0.1%) |
| Epilepsy | 92 (0.1%) |
| Mental or behavioural disorders | 87 (0.1%) |
| Hepatitis E | 83 (0.1%) |
| Fatty liver | 72 (0.1%) |
| Systemic lupus erythematosus | 69 (0.1%) |
| Thyroid nodule | 64 (0.1%) |
| Hepatic haemangioma | 58 (0.05%) |
| Pulmonary arterial hypertension | 35 (0.03%) |

**Supplementary Table 2. Full list of statistics on obstetric-relevant diagnostic indicators.**

| Obstetric-relevant indicators | Mean (SD) or Count (%) |
|---|---|
| **Gravidity** | 2.1 (1.2) |
| **Parity** | 1.5 (0.6) |
| **Abortion(s) or miscarriage(s)** | |
| 0 | 69,065 (58.2%) |
| 1 to 2 | 44,282 (37.3%) |
| 3 to 5 | 5,173 (4.4%) |
| above 5 | 169 (0.1%) |
| **Live birth(s)** | |
| 0 | 66,803 (56.3%) |
| 1 | 48,994 (41.3%) |
| 2 | 2,707 (2.3%) |
| 3 or above | 186 (0.2%) |
| **Spontaneous singleton delivery** | 62,818 (52.9%) |
| **Caesarean singleton delivery** | 51,506 (43.4%) |
| **Stillbirth** | 1,605 (1.4%) |
| **Gestational days** | 271 (18) |
| **Preterm birth** | 12,386 (10.4%) |
| **Singleton preterm birth** | 11,385 (9.6%) |
| **Twins or multiplets** | 4,926 (4.2%) |
| **Spontaneous preterm birth** | 6,738 (5.7%) |
| **Singleton spontaneous preterm birth** | 5,332 (4.5%) |
| **Singleton medical indicated preterm birth** | 6,053 (5.1%) |
| **Menarche age** (years) | 13.9 (1.4) |
| **Gestational weight gain** (kg) | 13.5 (6.0) |
| **Gestational diabetes mellitus** | 22,393 (18.9%) |
| **Gestational hypertension** | 2,465 (2.1%) |
| **Pre-eclampsia** | 6,553 (5.5%) |
| **Severe pre-eclampsia** | 4,342 (3.7%) |
| **Birth weight, singleton** (g) | 3,195 (580) |
| **Low birth weight infants, singleton** | 8,365 (7.0%) |
| **Apgar score, singleton, 1 min** | 9.7 (1.3) |
| **Apgar score, singleton, 5 min** | 9.9 (0.5) |
| **Neonatal sex, singleton** | |
| Male | 61,322 (51.7%) |
| Female | 57,289 (48.3%) |
| **First-degree obstetric lacerations** | 30,493 (25.7%) |
| **Second-degree obstetric lacerations** | 131 (0.1%) |
| **Third-degree obstetric lacerations** | 86 (0.1%) |
| **Intrauterine foetal distress** | 27,057 (22.8%) |
| **Premature rupture of membranes** | 25,160 (21.2%) |
| **Caesarean scar** | 24,587 (20.7%) |
| **Postpartum anaemia** | 21,787 (18.4%) |
| **Precipitate labour** | 8,962 (7.6%) |
| **Battledore placenta** | 8,921 (7.5%) |
| **Oligohydramnios** | 8,559 (7.2%) |
| **Macrosomia** | 7,712 (6.5%) |
| **Breech presentation** | 7,133 (6.0%) |
| **Postpartum haemorrhage** | 6,163 (5.2%) |
| **Intrauterine infection during pregnancy** | 5,337 (4.5%) |
| **Meconium-stained amniotic fluid** | 4,744 (4.0%) |
| **Intrahepatic cholestasis of pregnancy** | 4,188 (3.5%) |
| **Adherent placentas at delivery** | 3,719 (3.1%) |
| **Placenta previa** | 3,121 (2.6%) |
| **Placental abruption** | 2,988 (2.5%) |
| **Foetal growth restriction** | 2,447 (2.1%) |
| **Polyhydramnios** | 2,212 (1.9%) |
| **Foetal malformation** | 1,431 (1.2%) |
| **Vaginal birth after caesarean** | 984 (0.8%) |
| **Hypothyroidism in pregnancy** | 401 (0.3%) |
| **Foetal chromosome abnormalities** | 316 (0.3%) |
| **Twin pregnancies with single foetal death** | 266 (0.2%) |
| **Multifetal pregnancy reduction** | 253 (0.2%) |
| **Threatened uterine rupture** | 243 (0.2%) |
| **Foetal arrhythmia** | 230 (0.2%) |

**Supplementary Table 3. Full list of biochemical indices at parturient period and oral glucose tolerance test (OGTT) in second trimester, 24–28th gestational week.**

| Biochemical indices | Ref. range | Parturient period | Second trimester |
|---|---|---|---|
| **Fasting plasma glucose** (mmol/L) | ≤5.1 | - | 4.9 (1.2) |
| **OGTT 1h plasma glucose** (mmol/L) | ≤10 | - | 7.5 (2.1) |
| **OGTT 2h plasma glucose** (mmol/L) | ≤8.5 | - | 7.3 (1.6) |
| **White blood cell count** ($\times10^9$/L) | 3.5–9.5 | 10.7 (3.4) | 9.6 (2.2) |
| **Haemoglobin** (g/L) | 115–150 | 115.1 (13.8) | 114.7 (8.9) |
| **Platelet** ($\times10^9$/L) | 125–350 | 187.0 (49.1) | 209.9 (49.2) |
| **Neutrophils** (%) | 40.0–75.0 | 77.4 (6.9) | 74.9 (4.8) |
| **Lymphocytes** (%) | 20.0–50.0 | 15.4 (5.9) | 17.9 (4.2) |
| **Monocytes** (%) | 3.0–10.0 | 6.5 (1.7) | 6.0 (1.3) |
| **Eosinophils** (%) | 0.4–8.0 | 0.6 (0.6) | 1.0 (0.9) |
| **Basophils** (%) | ≤1.0 | 0.2 (0.1) | 0.3 (0.2) |
| **Haematocrit** (%) | 35.0–45.0 | 34.6 (34.7) | 34.3 (2.5) |
| **Serum protein, total** (g/L) | 65–85 | 59.3 (6.1) | 64.1 (3.6) |
| **Albumin** (g/L) | 40–55 | 33.1 (3.4) | 36.4 (2.2) |
| **Alanine aminotransferase** (U/L) | 7–40 | 15.2 (24.7) | 17.7 (17.4) |
| **Aspartate aminotransferase** (U/L) | 13–35 | 21.7 (18.3) | 18.7 (11.1) |
| **Bilirubin, total** (μmol/L) | ≤23.0 | 8.3 (4.0) | 6.9 (2.7) |
| **Bilirubin, conjugated** (μmol/L) | ≤4.0 | 2.6 (1.6) | 2.2 (1.0) |
| **Bilirubin, unconjugated** (μmol/L) | ≤19.0 | 5.8 (3.0) | 4.7 (2.1) |
| **Bile acids, total** (μmol/L) | 0–13 | 4.5 (6.7) | 2.3 (3.0) |
| **Lactate dehydrogenase** (U/L) | 120–250 | 208.6 (66.7) | 157.8 (29.9) |
| **Alkaline phosphatase** (U/L) | 35–100 | 155.6 (65.1) | 62.9 (20.0) |
| **5'-Nucleotidase** (U/L) | ≤10 | 4.9 (17.1) | 4.8 (2.0) |
| **Adenosine deaminase** (U/L) | ≤20 | 6.6 (1.8) | 5.9 (1.2) |
| **Gamma-glutamyl transpeptidase** (U/L) | 120–250 | 12.6 (11.9) | 12.2 (8.6) |
| **α-hydroxybutyrate dehydrogenase** (U/L) | 90–180 | 153.2 (38.2) | 117.6 (26.9) |
| **Creatine Kinase** (U/L) | 40–200 | 124.8 (167.1) | 38.4 (21.2) |
| **Creatinine** (μmol/L) | 41–73 | 52.6 (11.4) | 51.5 (10.6) |
| **Urea nitrogen** (mmol/L) | 2.6–7.5 | 3.1 (1.0) | 2.8 (0.7) |
| **Uric acid** (μmol/L) | 155–357 | 315.1 (76.2) | 237.4 (49.3) |
| **Triglycerides** (mmol/L) | 0.56–1.70 | 3.20 (1.60) | 2.40 (0.90) |
| **Cholesterol, total** (mmol/L) | 2.84–5.69 | 6.10 (1.30) | 6.10 (1.10) |
| **HDL-cholesterol** (mmol/L) | 1.03–1.55 | 1.70 (0.40) | 1.90 (0.40) |
| **LDL-cholesterol** (mmol/L) | 1.55–3.36 | 3.20 (1.00) | 3.10 (0.90) |
| **Potassium** (mmol/L) | 3.5–5.3 | 3.9 (0.3) | 4.0 (0.3) |
| **Sodium** (mmol/L) | 137.0–147.0 | 137.4 (2.2) | 137.7 (1.8) |
| **Chlorine** (mmol/L) | 99.0–110.0 | 105.1 (2.1) | 103.8 (2.1) |
| **Calcium, total** (mmol/L) | 2.11–2.52 | 2.20 (0.10) | 2.20 (0.10) |
| **Magnesium** (mmol/L) | 0.75–1.02 | 0.80 (0.10) | 0.80 (0.20) |
| **Phosphate** (mmol/L) | 0.85–1.51 | 1.20 (0.20) | 1.20 (0.20) |
| **Total iron binding capacity** (μmol/L) | 7.8–32.2 | 14.7 (8.3) | 17.3 (10.7) |
| **Hypersensitive C-reactive protein** (mg/L) | ≤5.0 | 17.7 (24.7) | 5.2 (6.5) |
| **Prealbumin** (g/L) | 180–350 | 203.4 (40.9) | 221.6 (30.2) |
| **Transferrin** (μmol/L) | 2.00–3.60 | 3.6 (0.7) | 3.6 (0.7) |
| **Glycosylated albumin** (%) | 10.8–17.1 | 11.6 (1.3) | 12.1 (0.9) |
| **Homocysteine** (μmol/L) | 5–15 | 7.9 (2.4) | 6.8 (2.1) |
| **Plasma glucose** (mmol/L) | ≤5.1 | 0.3 (1.5) | 1.5 (2.9) |
| **Cholylglycine** (μg/dL) | ≤270 | 232.5 (270.3) | 148.2 (134.5) |
| **Prothrombin time** (sec) | 11.8–13.9 | 12.6 (0.7) | 12.6 (0.5) |
| **International normalised ratio** | 0.89–1.90 | 0.98 (0.07) | 0.98 (0.05) |
| **Partial thromboplastin time, activated** (sec) | 29.7–41.8 | 33.2 (3.0) | 31.8 (2.3) |
| **Thrombin time** (sec) | 15.0–18.0 | 15.1 (1.0) | 14.9 (0.7) |
| **Fibrinogen** (g/L) | 1.90–3.80 | 4.6 (0.8) | 4.3 (0.7) |
| **Urine specific gravity** | 1.003–1.030 | 1.014 (0.007) | 1.014 (0.007) |
| **Proteinuria** | | | |
| - (Negative) | - | 113,724 (95.8%) | 63,600 (99.6%) |
| + | - | 3,328 (2.8%) | 206 (0.3%) |
| ++ | - | 1,226 (1.0%) | 51 (0.1%) |
| +++ | - | 411 (0.3%) | 30 (0.05%) |
| **Microscopic haematuria** | | | |
| - | - | 60,404 (50.9%) | 60,577 (94.8%) |
| + | - | 6,865 (5.8%) | 1,966 (3.1%) |
| ++ | - | 9,143 (7.7%) | 799 (1.3%) |
| +++ | - | 42,278 (35.6%) | 545 (0.9%) |
| **Glycosuria** | | | |
| - | - | 111,848 (94.2%) | 60,397 (94.5%) |
| + | - | 2,545 (2.1%) | 1,267 (2.0%) |
| ++ | - | 2,810 (2.4%) | 1,362 (2.1%) |
| +++ | - | 1,365 (1.1%) | 828 (1.3%) |
| ++++ | - | 122 (0.1%) | 21 (0.1%) |
| **Urine bilirubin** | | | |

| Biochemical indices | Ref. range | Parturient period | Second trimester |
|---|---|---|---|
| - | - | 117,884 (99.3%) | 63,747 (99.8%) |
| + | - | 707 (0.6%) | 106 (0.2%) |
| ++ | - | 63 (0.1%) | 26 (0.04%) |
| +++ | - | 34 (0.03%) | 6 (0.01%) |
| **Urine leucocyte** | | | |
| - | - | 85,636 (72.2%) | 39,018 (61.1%) |
| + | - | 11,060 (9.3%) | 11,769 (18.4%) |
| ++ | - | 8,947 (7.5%) | 5,392 (8.4%) |
| +++ | - | 13,047 (11.0%) | 7,708 (12.1%) |
| **Protein excretion, 24-hr** (g) | <0.15 | 1.10 (2.30) | 0.10 (0.10) |
